# Mathematical Analysis, Model and Prediction of COVID-19 Data

**DOI:** 10.1101/2020.08.04.20168195

**Authors:** Yit C. Tong

## Abstract

A simple and effective mathematical procedure for the description of observed COVID-19 data and calculation of future projections is presented. An exponential function E(t) with a time-varying Growth Constant k(t) is used. E(t) closely approximates observed COVID-19 Daily Confirmed Cases with NRMSD’s of 1 to 2%. An example of prediction of future cases is presented. The Effective Growth Rates of a discrete SIR model were estimated on the basis of k(t) for COVID-19 data for Germany, and were found to be consistent with those reported in a previous study (*1)*. The proposed procedure, which involves less than ten basic algebraic, logarithm and exponentiation operations for each data point, is suitable for use in promoting interdisciplinary research, exchange and sharing of information.

## Text

Mathematical analysis of empirical data provides insight into the structure and features of the underlying physical mechanism, and places constraints on the development and interpretation of mathematical models (*2,3*). In this study, mathematical curve fitting is applied to daily COVID-19 Confirmed Cases. The relationships between the time-varying parameter of the mathematical function used in the curve fitting and those of traditional SIR models are considered.

The mathematical presentation is kept at the level of elementary algebra. The computation for each daily data point involves a total of less than ten basic algebraic, logarithm and exponentiation operations (*4*). The proposed procedure is therefore suitable for use in promoting interdisciplinary research, exchange and sharing of information.

In this study, the observed 7-day moving average of COVID-19 daily Confirmed Cases, C(t), is represented by a time series sampled from an exponential function, E(t), with time-varying Growth Constant k(t) and intercept L(t),

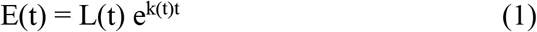

t is measured in days. 7-day averaging spans over the period from (t-3) to (t+3). For brevity, the discrete time series sampled from E(t) will also be referred to as E(t) in the following descriptions.

Although k(t) and L(t) can be determined by standard regression procedures, a simpler procedure that uses a numerically efficient algorithm for calculating E(t) that gives an adequate representation of C(t) is proposed. The algorithm also provides a simple means for the calculation of Projected Cases of E(t) for hypothetical scenarios of k(t) beyond the observation period of C(t) proposed by investigators.

In the algorithm, k(t) is chosen to be the approximate gradient of the natural logarithm of three successive observed data samples, C(t-1), C(t) and C(t+1),

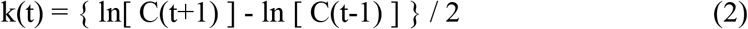

That is, E(t) is considered quasi-static with the same k(t) over the three samples.

Since the growth and decay of E(t) is governed by k(t), the time series E(t) should be adequately characterized by k(t) together with a single E(T), the value of E(t) on the T^th^ day. In other words, it should be possible to reconstruct an adequate approximation to the entire time series E(t) using k(t) together with a single initial E(T). In computational terms, the reconstruction starts at E(T) at time T, and successive E(t)’s at t before and after T are then calculated in accordance with (and guided by) k(t).

Applying Eqs. 1 and 2 to describe the time series of E(t) characterized by the same k(t) over three successive days, the following algorithm for the reconstruction is used,

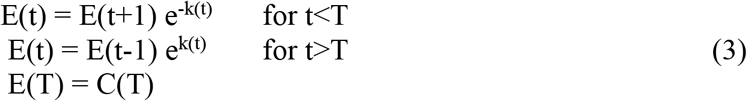

Two different formulas are used, one for forward difference (t<T) and the other backward difference (t>T), with an initial starting point at T. The purpose of using two formulas is purely computational, and Eq. 3 does not deviate from the mathematical relationships described in Eqs. 1 and 2.

Using Eq.3, calculation of Projected Cases of E(t) for hypothetical scenarios of k(t) beyond the observation period of C(t) is simply a continuation of the reconstruction process with its origin at T.

In this study, T is chosen to be the time point when the value of C(T) is the maximum of all observed C(t)’s. The maximum is chosen because the trend of observed C(t) and the reconstructed E(t) away from T would in general be more likely downhill than uphill. This will result in e^k(t)^, the scale factor, more likely to be less than one than greater than one. A scale factor that is less than one attenuates the propagation of errors between successive computational steps described in Eq. 3. These errors are introduced by the mismatch between C(t) and E(t).

The efficacy of this intuitive choice of T at maximum C(t) is supported quantitatively by comparing the mean squared errors (MSE’s) between C(t) and E(t) for all T’s within the observation period. The MSE’s for choosing C(T) in the vicinity of the maximum are consistently lower than the rest. The normalized root-mean-square deviation (NRMSD) is always within the range of 1 to 2%using this choice of the maximum, for COVID-19 data analyzed for five countries (*4*) showing a variety of trends in C(t).

For future data with very complex trends of C(t) such as those exhibiting multiple local maxima, several T’s can be used to reconstruct different sections of E(t) to achieve better approximation to C(t). Alternatively, for the purpose of more accurate future projections, the local maximum closest to the end of the observation period of C(t) can be used.

As an illustrative example, the mathematical procedure described in Eqs. 1-3 were applied to the 7-day averaged Confirmed Cases reported in the Worldometers Website (*5*) for Germany from 26-Feb to 3-July. The Worldometers data, apart from minor differences, are very close to those reported by WHO and Johns Hopkins, and were chosen for the present illustration because of its ease of accessibility. A spreadsheet detailing the complete series of observed 7-day averaged Confirmed Cases used for this example and implementation of Eqs. 2 and 3 can be found in (*4*).

Figure 1 shows the daily Observed Confirmed Cases C(t) for Germany as filled triangles, the Reconstructed E(t) as a solid line, and the estimated Growth Constant k(t) as a dotted line. T was chosen to be 31-Mar when the maximum of C(t) = 5837 occurred. The major ordinate on the left represents the values of C(t) and E(t), while the secondary ordinate on the right represents k(t). The vertical dotted line at 3-Jul indicates the end of observed data and the beginning of future projections.

Projections of E(t) for 20 days after 3-Jul were calculated as a continuation of the reconstruction defined by Eq. 3 initiated at time T (31-Mar) for an artificially generated sequence of k(t) that increased linearly from -0.012 to 0.2. Note that k(t) estimated using observed data at 3-Jul was -0.012. This is a hypothetical scenario where social distancing mitigation is gradually relaxed that would cause an increase in cases. Other scenarios for decrease or very little change in k(t) can also be easily explored. Scenarios chosen for study are community-specific, and can be formulated on the basis of trends of k(t) for data observed during different phases of the pandemic which are influenced by mitigation measures.

From Fig. 1, it can be seen that E(t) provides an excellent match to C(t), with a normalized root-mean-square deviation (NRMSD) of 1.24%.

With this close match, the characteristics of C(t) will be understood to be adequately represented by those of E(t) in the ensuing discussions.

**Fig. 1.**
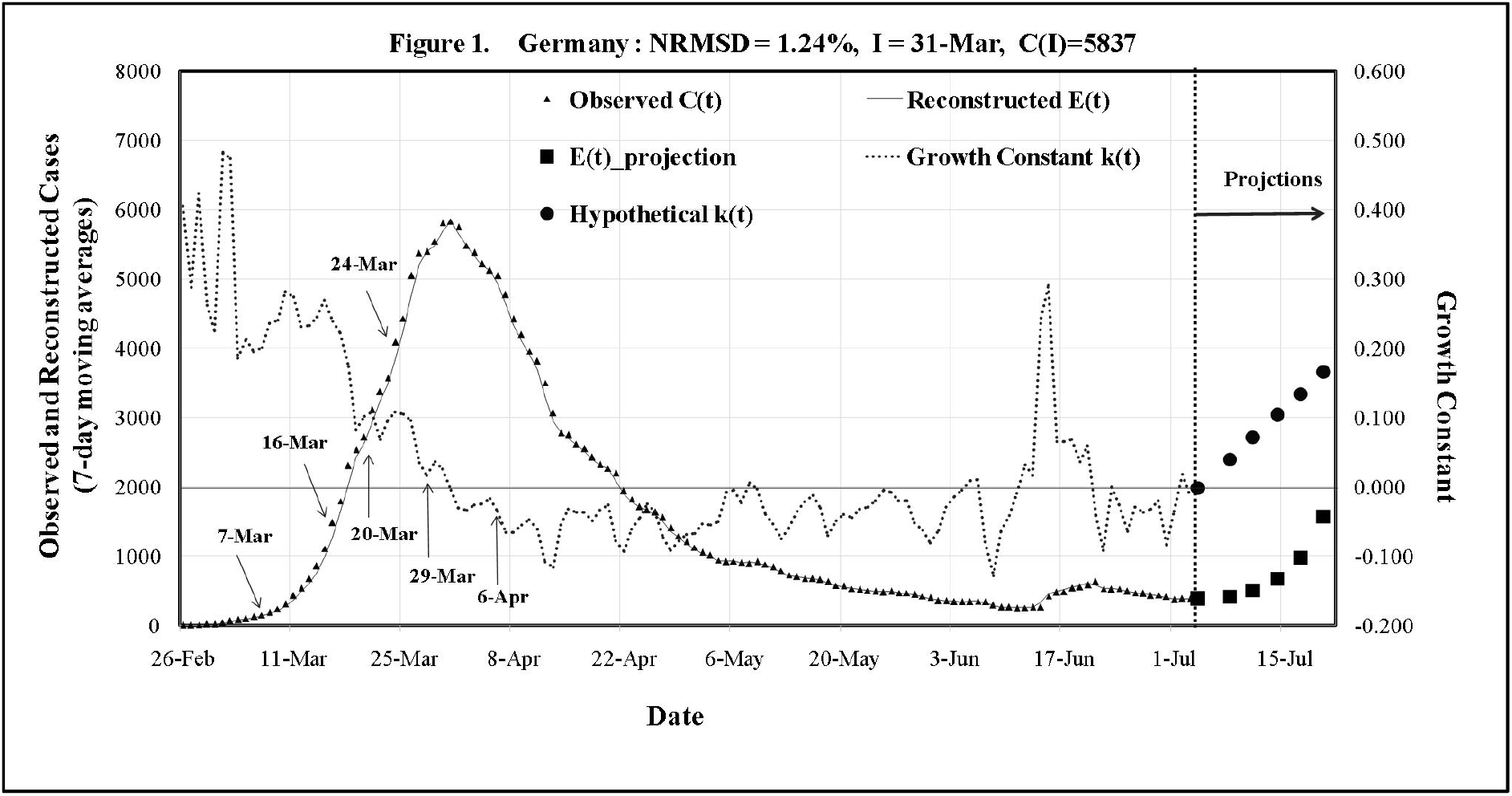
Daily Observed Confirmed Cases C(t), Reconstructed E(t) and Estimated Growth Constant k(t) for Germany: NRMSD = 1.24%, T = 31-Mar. Reconstruction of E(t) in accordance with Eq.3 was initiated at T = 31-Mar, when the maximum of C(t) occurred. E(t) provides an excellent match to observed C(t) with NRMSD = 1.24% for the Germany data. The vertical dotted line at 3-Jul indicates the end of observed data and the beginning of future projections. Projections of E(t) for 20 days after 3-Jul were calculated as a continuation of the reconstruction initiated at 31-Mar for an artificially generated increasing sequence of Hypothetical k(t) that increased linearly from -0.012 to 0.2. Note that k(t) estimated using observed data at 3-Jul was -0.012. The tilted arrows indicate the three dates of sequential interventions imposed by the German government. The vertical arrows indicate the matching sequence of dates 13 days after intervention. The values of k(t), which are close approximations to the Effective Growth Rates, at the three dates indicated by the vertical arrows are consistent with the corresponding Effective Growth Rates that were found to correlate well with the interventions reported in (*1*) for the Germany data.

As expected and by design, k(t) is consistent with the growth and decay pattern of E(t) and C(t), during the observation period that ended 3-Jul. k(t) has an initially large value of 0.269 at 15-Mar corresponding to the initial steep increase in E(t), reaches approximately zero when E(t) is maximum at 31-Mar, decreases to below zero corresponding to the continual decrease in E(t), and hovers within a narrow range close to zero of mostly negative or occasionally positive values corresponding to an initially declining and then a mostly level E(t) until 3-Jul. In mathematical terms, from Eq. 3, this is understandable in that E(t) grows for k(t)>0 (e^k(t)^>1), no change when k(t)=0 (e^k(t)^=1), and decays when k(t)<0 (e^k(t)^<1). Note that faster growth or decay would result for larger absolute values of k(t), and slower for smaller absolute k(t).

A Growth Factor over a specified number of days can be calculated to reflect the degree of growth or decay when k is a constant. For a given k, the daily scale factor is r=e^k^. If we consider a geometric series of N+1 days: {E(t), rE(t), r^2^E(t),……., r^N^E(t)}, E(t) would grow by a factor of G = r^N^ in N days, resulting in the following relationship,

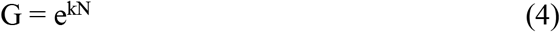

For example, for k = 0.087 and N = 8, G is equal to 2, corresponding to a doubling of E(t) in 8 days. In general, Eq. 4 can be used to calculate G, k or N for any combination of the other two variables. Eq. 4 provides a rule-of-thumb means for accessing the consequences of various scenarios defined by hypothetical k(t)’s.

In terms of physical modeling, the following conventional discrete SIR model (1,6,7) is considered,

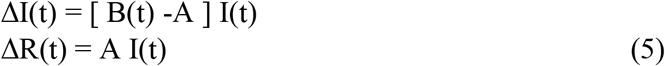

The Infectious Compartment, I(t), models the number of currently infectious individuals in the community, who are capable of infecting other susceptible members. The Removal Compartment, R(t), models individuals removed from I and become no longer infectious. B(t)is the Spreading Rate, while A is the Removal Rate. On day t, the daily input to the I Compartment is B(t)I(t), the number of newly infected, while the daily output from I is AI(t), the number removed from I and migrated to Compartment R. [B(t)-A] is the Effective Growth Rate.

Note that in most SIR models, 1/A is a constant and taken to be the Infectious Period, the number of days over which any member of the I compartment can be considered infectious before being removed. ΔI(t) and ΔR(t) represent the daily changes of the I and R Compartments, respectively. I(t) increases when B(t) > A, no change when B(t) = A, and decreases when B(t) < A.

If E(t) is considered to be equivalent to ΔR(t), the daily change of R, k(t) in Eq. 1 and Fig. 1 for E(t) is also the Growth Constant for I(t). This is because, from Eq. 1 and by equating ΔR(t) to E(t),

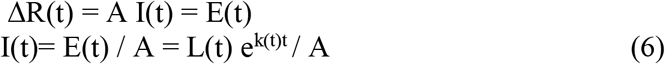

For Simple SIR models, I(t) is therefore a linearly scaled up version of E(t) by a constant factor 1/A as described in Eq. 6. For this reason and brevity, I(t) is not included in the illustration in Fig. 1 for the Germany data.

It is reasonable to consider E(t), the Reconstructed Confirmed Cases, to be equivalent to ΔR(t), individuals who migrate daily from the I Compartment to the R Compartment after an Infectious Period of 1/A days, because they will be either quarantined, hospitalized, deceased or immunized, and would cease to infect other members of the community.

As far as parameter estimation of SIR models is concerned, the Effective Growth Rate [B(t)-A] can be estimated on the basis of k(t). Noting that from Eqs. 3 and 6, the daily scale factor for I(t) is e^k(t)^. This leads to,

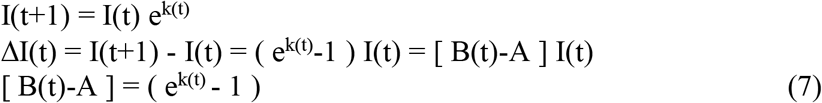

Note that [B(t)-A] = (e^k(t)^-1) is approximately equal to k(t) for small |k(t)|<0.1. For example, for k(t)=0.100, [B(t)-A] = (e^k(t)^-1) = 0.105.

Since k(t) is the same for both E(t) and I(t), Eq. 4 for E(t) is also applicable to I(t). For a given constant k, the daily scale factor for I(t) is also e^k^. If we consider a geometric series of N+1 days: {I(t), rI(t), r^2^I(t), r^N^I(t)}, I(t) would grow by a factor of G = r^N^in N days. Renaming G as G’ for SIR models, and equating N to 1/A,

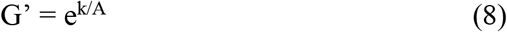

It can be seen from the above geometric series, the difference between two successive terms, r^n^I(t) and r^n-1^I(t), is (r-1) times r^n-1^I(t43). This is consistent with Eq. 7 because r=e^k^.

G’ in Eq. 8 can be considered to be equivalent to the Effective Reproduction Number in SIR Models, which gives an indication of the number of new infections generated over the Infectious Period by each infectious before its migration from I to R. Using the same numerical example of Eq. 4, for k = 0.087 and Infectious Period 1/A = 8, G’ is equal to 2, corresponding to a doubling of I(t), or 2 new infections generated by each infectious over the Infectious Period.

The SIR Effective Growth Rates [B(t)-A] = (e^k(t)^ - 1) estimated by Eq. 7 for the Germany data shown in Fig. 1 are consistent with those reported in (*1*) estimated using a different method for the same data. In (*1*), the Effective Growth Rates for three change points spanning 17 days were reported as a decreasing sequence of 0.12, 0.02 and -0.03 corresponding to three government interventions on 7-Mar, 16-Mar and 24-Mar, respectively. The three dates are indicated by the tilted arrows in Fig. 1. Note that the second intervention had been imposed 9 days after the first while the third was imposed 8 days after the second. In Fig. 1, consider the decreasing sequence of 0.102, 0.036 and -0.035 of k(t) ≈ [B(t)-A] at 20-Mar, 29-Mar and 6-Apr, respectively. These three dates are indicated by the vertical arrows in Fig. 1. The two sequences of values of Effective Growth Rate can be considered very similar given the nature of the available observed data, and the difference between the two estimation methods used in (*1*) and the present study. The two corresponding sequences of dates have the same two intervals of separation, 9 days and 8 days. The difference in absolute dates of 13 days (between 7-Mar and 20-Mar) can be attributed to delays involved in the time required to achieve the desired effectiveness of each intervention (e.g. 5 days), and time shift due to averaging and reporting (e.g. 8 days).

Although [B(t)-A)] can be estimated by (e^k(t)^-1) using Eq. 7, there is no unique solution for either B(t) or A, using the method of analysis in this study. In order to complete the parameter estimation, other consideration and assumptions can be made to propose a suitable A. For example, 1/A can be taken to be approximately 8 days (Removal Rate of A=0.125) for the Germany data in accordance with community-specific information provided in (*1*). In (*1*), 8 days was proposed to be the typical reporting/detection delay (before removal) caused by incubation, symptoms development and testing. If we assume that this delay of 8 days is the Infectious Period 1/A, then, using our previous numerical example, k=0.087 and A=1/8=0.125 would result in (e^k(t)^-1)=0.091 and B=0.216. It can be easily shown that applying B=0.216 and A=0.125 to Eq. 7 to calculate ΔI(t) = I(t+1)-I(t) = [B(t)-A]I(t) recursively for 8 successive days, a doubling of I(t) would occur, corresponding to an Effective Reproduction Number of 2.

Finally, the number of daily Death Cases, D(t), is of particular interest in the management of pandemics. The Death statistics is time-varying and follows a downward trend because of progressive improvements in clinical care. A Death-to-Confirmed-Ratio can be approximated by the average ratio of Death to Confirmed Cases in the last ten days of the observation period. For communities who are in the early stage of the pandemic, a time delayed time series of Confirmed Cases C(t-τ), to correct for the time delay τ between the date of Confirmation and date of Death, can be used in estimating the ratio. Projected daily Death Cases can then be calculated by multiplying the Projected Confirmed Cases E(t) in Fig. 1 by the estimated Death-to-Confirmed-Ratio.

## Data Availability

Public domain from Worldometers, Johns Hopkins or WHO

https://www.worldometers.info/coronavirus

https://coronavirus.jhu.edu/map

https://covid19.who.int/

